# Racial and Ethnic Inequities in Wealth and Health: Evidence from a Multiethnic Survey in NYC

**DOI:** 10.64898/2026.02.09.26345760

**Authors:** Judy Fordjuoh, Sarah J. Bloomstone, Yaoyu Zhong, Shadi Chamany, Ellen Wiewel, Duncan Maru, Amaka Anekwe, Luisa N. Borrell, Mustafa Hussein, Zachary Shahn, Trenton M. White, Ayman El-Mohandes, William Darity, Michelle Morse

**Author notes:** **Corresponding Author Information:** Judy Fordjuoh New York City Department of Health 42-09 28^th^ St, desk 11-13, Long Island City, NY 11101 /.

## Abstract

**Objective:** To examine racial and ethnic inequities in wealth and health among New York City adults.

**Methods:** We conducted the 2024 NYC Racial Wealth and Health Gap Survey using a stratified quota sample of 2,866 adults across 11 racial and ethnic groups. Wealth was measured through self-reported assets and debts, and health through self-reported status and psychological distress. We calculated descriptive statistics across groups and used quantile regression to test for significant differences in assets and debts compared with White respondents.

**Results:** White and Chinese respondents reported the highest median net worth ($142,000 and $320,000), while Other Black and Puerto Rican respondents reported the lowest ($25 and $160). Lower wealth was associated with poorer health and higher psychological distress. Prevalence of excellent or very good health increased from 36% in the lowest wealth quartile to 59% in the highest, with the steepest wealth–health gradients among Chinese and Multiracial respondents.

**Conclusion:** Wealth inequities are linked to health disparities across racial and ethnic groups in New York City. Surveillance of local wealth data can guide equity-focused policies addressing economic and racial drivers of health disparities.

## Introduction

Wealth is often measured as net worth at a point in time, or the difference between the value of assets owned and the amount owed in debts [1]. Wealth inequality in the United States is significant. A national survey conducted in 2022 revealed that the median net worth of non-Hispanic white families was six times that of non-Hispanic Black families, and five times that of Hispanic/Latino families [1]. When data disaggregated by racial and ethnic groups are available, even starker gaps emerge. In Boston, for example, non-Hispanic white households had a self-reported net worth of $245,000, while US-born Black and Dominican households each had a net worth of less than $10 [2].

Racial and ethnic wealth gaps have persisted for generations. In 1950, the white-to-Black per capita wealth ratio was approximately 7 to 1, and by 2020, it remained largely unchanged at 6 to 1, meaning that little progress has been made in closing the racial wealth gap over the past seven decades [3]. When measured as the monetary difference between white and Black wealth adjusted for inflation, the gap increased 18% from $981,170 in 2016 to $1.155 million in 2022 (in 2022 dollars), even as the white-to-Black wealth ratio fell slightly, from 6.76 to 6.47, during that period [4, 5].

Governmental policies at the federal, state, and local levels have long facilitated wealth-building and asset accumulation for white Americans while denying the same opportunities to other racial and ethnic groups. Although inequities in wealth reflect the “intersection of the cumulative impact of United States racial history and the racially uneven transmission of resources across generations, the specific origins of these inequities vary across groups” [4, 6]. For example, the wealth gap between Indigenous Americans and white Americans is largely the result of the dispossession and occupation of native lands, [7] through policies such as the Indian Removal Act and the Dawes Act. The Black-white wealth gap reflects the legacies of enslavement, discrimination, and government policies like the Homestead Act of 1862, seizure of Black properties via 100 massacres between the Civil War and World War II, redlining, [8] the GI Bill, urban renewal, and the federal highway program. Collectively, these policies established financial systems that extract wealth and hinder its intergenerational transfer [4, 9–11].

Direct transfers of wealth, such as inheritances, gifts of property, or in-kind transfers, including access to quality education and high-amenity neighborhoods, play a decisive role in shaping a person’s wealth trajectory and mobility [9, 12, 13]. Wealth empowers individuals and households to save money, invest in health-promoting behaviors, weather financial shocks, and pay for housing, education, and healthcare without incurring short- or long-term debt.

A growing body of evidence demonstrates that net worth is associated with a variety of health indicators including self-rated health, [14, 15] chronic conditions, [16] depression, [17] mental health, [15] mortality, [18] and functional impairment [19]. However, this relationship is nuanced, as net worth and its components relate to health in different and complex ways. Specific assets such as savings accounts, stock ownership, and homeownership are independently associated with better health, while specific debts such as student loans, credit card debt, and medical debt are independently associated with worse health [20]. Several modeling studies demonstrate how eliminating wealth inequality can reduce health inequities, including COVID-19 transmission and gaps in longevity [21–23].

National surveys, such as the Panel Study of Income Dynamics and the Survey of Income and Program Participation, offer valuable information about income, wealth, and in some cases the relationship between wealth and health, but they are unable to capture the full scope of inequities in wealth distribution or their connection to health at the local level. These surveys rely on broad racial and ethnic categories that can obscure meaningful differences in lived experiences.

The Color of Wealth study series has addressed some of these gaps by examining wealth at the local level in jurisdictions across the US and employing a sampling framework that allows the disaggregation of data by respondent’s country of origin or ancestry. These studies oversampled according to the specific racial and ethnic composition of each participating city and used the National Asset Scorecard for Communities of Color to measure respondents’ assets, debts, financial resources, savings, investments, and demographic characteristics. This approach revealed important differences in wealth across ancestry groups. However, the existing Color of Wealth studies were designed to examine local racial wealth inequality and did not incorporate health measures.

A study clarifying the relationship between wealth and health within disaggregated racial and ethnic groups has not been conducted in New York City. Investigating wealth and health in New York City is particularly important because despite being considered one of the wealthiest cities in the nation, it is also characterized by socioeconomic inequality, vast racial and ethnic diversity, and a complex mix of public and private insurance programs that differ from those in other cities in the U.S. New York City’s large immigrant population, varied labor markets, and dense healthcare infrastructure may amplify or mitigate inequities in distinct ways. These structural features shape access to care, affordability, and health outcomes in ways that are hard to infer from research conducted elsewhere. As a result, localized data are necessary to understand the specific dynamics of wealth and health in New York City.

To address this gap, the New York City Department of Health and Mental Hygiene (NYC Health Department), in collaboration with the City University of New York (CUNY) Graduate School of Public Health and Health Policy, developed the 2024 NYC Racial Wealth and Health Gap Survey. Building on a series of Color of Wealth studies [2, 24–28], this survey fills a critical gap in the NYC data landscape, by generating detailed and locally grounded evidence on wealth and health among residents, disaggregated by race, ethnicity, and ancestry. This research aimed to examine how wealth relates to key health status indicators across 11 racial and ethnic groups and to identify inequities that may not be observable using national datasets. By linking disaggregated wealth information with measures of health, the study provides actionable data to inform strategies intended to reduce inequities in both wealth and health.

## Methods

### Sample Selection/Study Population

To identify the most salient racial and ethnic groups and their subgroups in New York City, American Community Survey data was used to select 11 mutually exclusive racial and ethnic groups to survey: Puerto Rican, Dominican, or Other Hispanic ancestry; African American, Caribbean or West Indian, or Other Black ancestry; Chinese or Other Asian or Pacific Islander (API) ancestry; Indigenous ancestry, including American Indian, Native, First Nations, Indigenous Peoples of the Americas, or Alaska Native ancestries; White; and Multiracial or some other race, including Middle Eastern or North African. Some of these are the largest racial and ethnic groups, while others are smaller but have a notable presence in NYC and merit study of their wealth and health. We created strata based on project funding (capping at 3200 participants due to logistical constraints), reflecting either relatively larger or smaller population sizes. Therefore, the strata consist of eight ancestry subgroups with a target of 360 participants each, and three subgroups with a target between 125 and 200 participants each.

We partnered with Consensus Strategies, LLC (Consensus), a consulting group specializing in polling, survey research, and community advocacy. The study population was drawn using random quota-based sampling from two pre-existing Consensus databases. The first includes approximately 5.1 million NYC adults with email addresses, phone numbers, or both, constructed using Census, voter registration, and utility and credit card bill information. While it covers 79% of the overall adult population using 2021 American Community Survey 5-year estimates, it includes a smaller proportion of NYC’s adult Latino and Asian adult populations than other racial/ethnic groups (60% of Latinos and Asians, vs. ≥85% of other groups). The second is a database of 2.1 million panel members. Outreach began with text messages, followed by email and then panel outreach, until the pre-determined sample sizes for each of the 11 racial and ethnic ancestry groups were reached.

Stratified quota sampling in 100,000-person batches was done until we reached the target responses for each ancestry stratum, monitored through participant survey responses on race, ethnicity, and ancestry questions. We constructed racial and ethnic groups based on respondents’ self-reported races and first and second ancestry or ethnic origin, defined as the country or territory of origin for themselves, their parents, grandparents, or ancestors.

We recruited most participants through English-language invitations via text messages and emails. To reach groups with a lower representation in the population, such as indigenous and Black Latino New Yorkers, we utilized a separate panel of 116,000 people, also maintained by Consensus.

This study was approved by the Institutional Review Board (IRB) of Emerson College, the academic partner of Consensus (IRB protocol no. 24-015-F-E-12/1, [12/1/2023] [modified 5/29/2024]).

### Survey Design and Data Collection

We developed the NYC Racial Wealth and Health Gap survey instrument by adapting the National Asset Scorecard for Communities of Color (NASCC),[29] which was used in the Color of Wealth surveys across several major U.S. cities [2, 24–28]. Like the NASCC, our instrument measures specific assets, debts, financial resources, personal savings, investment activity, and other financial experiences, and collects demographic data such as race, ethnicity, ancestry, age, sex, educational attainment, household composition, nativity, income, and family background. Our instrument expands on the NASCC by including questions about physical and mental health status, as well as healthcare access and utilization.

We collected data in June 2024 using Qualtrics, a secure online survey platform. Participants opted to participate in the web-based survey and provided digital consent via a checkbox. The survey contained 232 questions and was initially estimated to take approximately 30-45 minutes to complete; in practice, respondents spent an average of two hours. Participants could pause or leave the website and return to complete the survey as many times as they needed within a 2- week period. The survey was conducted in English, Spanish, simplified and traditional Chinese, Bengali, Korean, and Kreyòl, as these are some of the most spoken and/or written languages in NYC [30]. Translations were completed by Consensus to ensure accuracy and cultural appropriateness. Individuals who completed the survey received a $25 gift card.

### Measures

All the measures in this study were self-reported. From the instrument, we constructed the 11 racial/ethnic groupings as listed above. To reflect the survey’s focus on ancestry, we applied a classification hierarchy in which respondents who identified as Hispanic or Latino were categorized into one of the Hispanic subgroups, regardless of their racial identities (e.g., Black, white, Indigenous).

We used household net worth as the measure for wealth, calculated as the difference between the value of all reported assets and debts. Assets included liquid assets (i.e., checking and savings accounts, stocks, mutual funds, investment trusts, individual retirement accounts, private annuities, and pension or retirement plans) and tangible assets (i.e., the value of primary residence, the value of other real estate, a business or farm, and the value of all vehicles owned). Debts included unsecured debts from credit cards, installment loans, student loans, medical bills, legal bills, or loans from friends or relatives, and secured debts, including mortgages and vehicle debt.

Measures of physical health included self-reported health status (excellent/very good, good, fair/poor), presence of specific chronic diseases (asthma, diabetes, and/or hypertension) as told by a doctor or other healthcare professional, and psychological distress. Psychological distress was measured using the 6-item Kessler scale and then dichotomized into low and high levels of distress using the standard threshold.

Of the 5,381 adults who began the NYC Racial Wealth and Health Gap Survey, 3,531 completed all required questions, yielding a 66% completion rate. Out of a total of 5,381, we excluded participants who: 1) did not complete the entire survey (n= 1847); 2) exhibited unrealistically low engagement (i.e., survey duration under 7 minutes; n=71); 3) had responses with validity issues, such as consistently entering sequential numbers (e.g., 1234) or repeating numbers (e.g., 1111; n=244); 4) reported having a specific asset or debt but failed to provide a corresponding value (n=337); 5) had outlier responses, such as individuals reporting assets or debts in the trillions, and those who entered negative values for pensions (n=5); and 6) had responses that did not allow for race or ethnicity to be determined (n=11). These exclusions yielded a final analytical sample of 2,866 participants.

### Data Analysis

We first estimated the median, first quartile (Q1), and third quartile (Q3) of wealth across 11 racial and ethnic groups, as well as across additional demographic characteristics such as age, gender, perceived racial bias or discrimination, place of birth, borough, education, employment status, household size, and risk of food insecurity. We further examined distributions of wealth quartiles, as well as distributions, medians, and corresponding 95% confidence intervals for detailed assets and debts as described in Measures section, across 11 racial and ethnic groups. We also tested whether the median of each wealth component differed significantly between White participants and each of the other racial/ethnic groups. To explore how wealth relates to health status, we examined distributions of wealth quartiles, as well as median, Q1, and Q3, by the three measures of physical health.

We used bivariable quantile regression to estimate median, Q1, and Q3, as well as pairwise differences in median of detailed asset and debt between each racial/ethnic group and the White reference group. Wealth data are highly skewed and often contain extreme values. Unlike simple descriptive procedures, quantile regression provides a regression-based approach that accommodates skewed wealth distributions, produces valid confidence intervals for medians, and enables direct comparison of median differences across groups. This approach allows us to rigorously quantify disparities in wealth and its components while accurately reflecting distributional differences across racial and ethnic groups.

All data management and analyses were conducted using SAS 9.4.

## Results

Of the analytic sample (n=2,866), 25% are aged 35-44, 57% are cisgender women, and 69% are employed. Half of the sample hold a bachelor’s degree or higher (51%), two-thirds rent their primary residence (66%), and three-quarters have 1-2 people in their household (75%; Table 1).

**Table 1.** Household Net Worth by Sample Demographics and Health Characteristics: Adults ages 18 and older in New York City, June 2024. Source: New York City Racial Wealth and Health Survey

|  | Total |  | Net Worth<sup>a</sup> |  |  |  |
| --- | --- | --- | --- | --- | --- | --- |
|  | N | Column % | Median amount in U.S. Dollars | Q1 in U.S. Dollars | Q3 in U.S. Dollars | Percent difference from the group with the highest median |
| **Overall Sample** | **2,866** | **100.0** |  |  |  |  |
| **Demographic Variables** |  |  |  |  |  |  |
| **Age** |  |  |  |  |  |  |
| 18-24 | 423 | 14.8 | 5,000 | 0 | 100,000 | -94 |
| 25-34 | 660 | 23.0 | 7,500 | 0 | 187,000 | -91 |
| 35-44 | 704 | 24.6 | 14,000 | 0 | 374,000 | -83 |
| 45-54 | 460 | 16.1 | 23,005 | 0 | 475,100 | -73 |
| 55-64 | 384 | 13.4 | 10,200 | 0 | 548,678 | -88 |
| 65+ | 235 | 8.2 | 84,000 | 0 | 1,260,000 | 0 |
| **Gender** |  |  |  |  |  |  |
| Cisgender man | 1,103 | 38.5 | 50,000 | 2 | 650,000 | 0 |
| Cisgender woman | 1,618 | 56.5 | 5,000 | 0 | 257,400 | -90 |
| Transgender, non-binary, or another gender identity not listed | 48 | 1.7 | 600 | 0 | 51,000 | -99 |
| Not sure or prefer not to answer | 97 | 3.4 | 0 | 0 | 5,000 | -100 |
| **Racial and Ethnic Groups** |  |  |  |  |  |  |
| **Latino** |  |  |  |  |  |  |
| Puerto Rican | 329 | 11.5 | 160 | 0 | 50,009 | -100 |
| Dominican | 287 | 10.0 | 300 | 0 | 35,000 | -100 |
| Other Latino | 354 | 12.4 | 5,000 | 0 | 200,000 | -98 |
| **Non-Latino** |  |  |  |  |  |  |
| **Black** |  |  |  |  |  |  |
| African American | 393 | 13.7 | 500 | 0 | 82,000 | -100 |
| Caribbean or West Indian | 246 | 8.6 | 8,800 | -900 | 372,150 | -97 |
| Other Black | 157 | 5.5 | 25 | 0 | 25,897 | -100 |
| **Asian or Pacific Islander** |  |  |  |  |  |  |
| Chinese | 252 | 8.8 | 320,000 | 22,000 | 1,080,000 | 0 |
| Other Asian or Pacific Islander | 277 | 9.7 | 107,000 | 1,850 | 754,000 | -67 |
| **American Indian, Native, First Nations, Indigenous Peoples of the Americas, or Alaska Native** | 23 | 0.8 | 1,000 | 0 | 48,000 | -100 |
| **Multi-race or some other race (including Middle Eastern/North African)** | 137 | 4.8 | 5,000 | -1,198 | 441,000 | -98 |
| **White** | 411 | 14.3 | 142,000 | 1,000 | 1,100,000 | -56 |
| **Perceived Racial Bias/Discrimination** |  |  |  |  |  |  |
| Worse than other races, ethnicities, or tribes | 621 | 21.7 | 2,480 | -5,000 | 267,000 | -97 |
| Same as than other races, ethnicities, or tribes | 1,517 | 52.9 | 17,000 | 0 | 393,425 | -80 |
| Better than people of other races, ethnicities, or tribes | 346 | 12.1 | 85,000 | 600 | 618,000 | 0 |
| Don't know | 382 | 13.3 | 150 | 0 | 150,000 | -100 |
| **Place of Birth** |  |  |  |  |  |  |
| In the United States, including U.S. territories | 2,167 | 75.6 | 10,880 | 0 | 350,000 | -28 |
| Outside of the United States | 699 | 24.4 | 15,100 | 0 | 410,000 | 0 |
| **Borough** |  |  |  |  |  |  |
| The Bronx | 618 | 21.6 | 210 | 0 | 61,000 | -100 |
| Brooklyn | 838 | 29.2 | 12,000 | 0 | 365,000 | -89 |
| Manhattan | 652 | 22.7 | 30,000 | 0 | 597,500 | -73 |
| Queens | 662 | 23.1 | 31,000 | 0 | 534,800 | -72 |
| Staten Island | 96 | 3.3 | 110,000 | 1 | 575,600 | 0 |
| **Education** |  |  |  |  |  |  |
| High school diploma or less | 864 | 30.1 | 50 | 0 | 20,000 | -100 |
| Trade or Vocational School, and Some College | 549 | 19.2 | 3,000 | 0 | 75,000 | -98 |
| Bachelor's Degree and higher | 1,448 | 50.5 | 132,000 | 2 | 822,000 | 0 |
| Don't know/missing | 5 | 0.2 | 600 | 0 | 5,002 | -100 |
| **Employment <sup>b</sup>** |  |  |  |  |  |  |
| Employed | 1,988 | 69.4 | 31,000 | 0 | 459,000 | 0 |
| Unemployed | 878 | 30.6 | 100 | 0 | 90,000 | -100 |
| **Employment Sector** |  |  |  |  |  |  |
| A private company | 1,176 | 41.0 | 29,000 | 0 | 459,000 | 0 |
| Family business | 114 | 4.0 | 9,720 | 0 | 510,000 | -66 |
| Non-profit | 378 | 13.2 | 20,160 | 0 | 380,000 | -30 |
| Local government | 322 | 11.2 | 26,002 | 0 | 504,600 | -10 |
| State government | 211 | 7.4 | 10,000 | 0 | 267,000 | -66 |
| Federal government | 82 | 2.9 | 4,024 | 0 | 375,000 | -86 |
| Independent contractor or<br>sole proprietor | 233 | 8.1 | 15,005 | 0 | 267,000 | -48 |
| Something else /Don't know | 292 | 10.2 | 0 | 0 | 28,300 | -100 |
| Never worked | 58 | 2.0 | 0 | 0 | 700 | -100 |
| **Marital Status** |  |  |  |  |  |  |
| Single and never married | 1,466 | 51.2 | 2,000 | 0 | 137,500 | -99 |
| Married | 756 | 26.4 | 200,000 | 2,000 | 1,011,700 | 0 |
| Living with a partner (but<br>not married) | 291 | 10.2 | 20,000 | 0 | 260,000 | -90 |
| Widowed | 75 | 2.6 | 10 | -1,500 | 115,000 | -100 |
| Divorced or separated | 278 | 9.7 | 200 | -4,700 | 212,120 | -100 |
| **Homeownership** |  |  |  |  |  |  |
| Owned (by you or someone<br>in your family unit) | 790 | 27.6 | 710,000 | 267,000 | 1,650,000 | 0 |
| Rented | 1,893 | 66.1 | 500 | -600 | 43,400 | -100 |
| Other/Unknown residence<br>status | 183 | 6.4 | 0 | 0 | 500 | -100 |
| **Household Size** |  |  |  |  |  |  |
| 1-2 people | 2,137 | 74.6 | 12,000 | 0 | 362,000 | -20 |
| 3-5 people | 651 | 22.7 | 15,005 | 0 | 365,000 | 0 |
| 6+ people | 53 | 1.8 | 10,100 | 6 | 601,800 | -33 |
| Did not respond to this<br>question | 25 | 0.9 | 0 | 0 | 1,000 | -100 |
| **Risk of Food Insecurity** |  |  |  |  |  |  |
| At risk of food insecurity | 1,291 | 45.1 | 100 | -1,198 | 32,456 | -100 |
| Not at risk of food<br>insecurity | 1,429 | 49.9 | 130,500 | 1,000 | 822,800 | 0 |
| Do Not Know | 146 | 5.1 | 0 | 0 | 60,000 | -100 |
| **Public Assistance** |  |  |  |  |  |  |
| Yes, my family members<br>and I receive public<br>assistance and/or income<br>support | 1,055 | 36.8 | 100 | 0 | 35,000 | -100 |
| No, my family members and<br>I do not receive/ I am<br>unsure if my family and I<br>receive public assistance | 1,811 | 63.2 | 54,000 | 0 | 635,186 | 0 |
| **Health status and Care Variables** |  |  |  |  |  |  |
| **Self-Reported Health** |  |  |  |  |  |  |
| Excellent/Very Good | 1,360 | 47.5 | 37,000 | 0 | 570,000 | 0 |
| Good | 967 | 33.7 | 8,800 | 0 | 281,500 | -76 |
| Fair/Poor | 539 | 18.8 | 100 | -4,000 | 66,500 | -100 |
| **Presence of a Chronic Disease (i.e. diabetes, hypertension, asthma)<sup>c</sup>** |  |  |  |  |  |  |
| No Chronic Disease | 1,536 | 53.6 | 15,200 | 0 | 400,800 | 0 |
| Chronic Disease | 1,330 | 46.4 | 10,000 | 0 | 317,000 | -34 |
| **Psychological Distress** |  |  |  |  |  |  |
| Low Psychological Distress or No Psychological Distress | 2,264 | 79.0 | 20,000 | 0 | 435,000 | 0 |
| High Psychological Distress | 602 | 21.0 | 2,000 | -1,500 | 105,000 | -90 |

<sup>a</sup> Net worth is the total wealth of the household, taking account of all financial assets owned minus the value of all outstanding debts or liabilities.
<sup>b</sup> Unemployment includes those not in the workforce, retired, currently in school and do not work, as well as people who maintain household for no pay
<sup>c</sup> Presence of a chronic disease does not include individuals with a chronic disease diagnosis only during pregnancy (e.g., diabetes or hypertension diagnosed only during pregnancy).
Note: To understand the degree and direction of bias within our sample, we compared the percentages of several sociodemographic variables such as race/ethnicity, gender identity, age, and employment, between our sample and New York City's population using 2023 American Community Survey data.

### Wealth by Racial/Ethnic Groups and Other Demographics

In each demographic characteristic of our sample, median net worth is highest among individuals who are aged 65+ ($84,000), identify as a cisgender man ($50,000), have a bachelor’s degree or higher ($132,000), are employed ($31,000), are homeowners ($710,000), are married ($200,000), or are foreign-born ($15,100) (Table 1).

Substantial gaps in net worth across racial and ethnic groups are present. Chinese respondents report the highest median net worth ($320,000), followed by white respondents ($142,000), while Puerto Rican ($160) and Other Black respondents have the lowest ($25) (Table 1). Except for White, Chinese, and API respondents, the bottom quartile of wealth among all other groups was either zero or negative.

Besides Caribbean or West Indian respondents, at least 55% of all remaining Black or Hispanic racial and ethnic groups are in the lowest two wealth quartiles. In contrast, more than 40% of White and Chinese respondents are in the highest wealth quartile (Supplementary Table 1).

### Net Worth Components by Racial/Ethnic Groups

Assets and debts are also unequally distributed across groups. The Chinese, White, and Other API groups report higher median values of tangible and liquid assets, contributing to greater total asset holdings ($480,000, $273,000, and $265,090, respectively), compared with Other Black, Puerto Rican, and Dominican groups, for example (total median asset accumulations of $11,000, $15,000, and $17,000, respectively) (Table 2). These differences are statistically significant, with Puerto Rican, Dominican, African American, Caribbean, and Other Black adults all reporting significantly lower total asset values compared with Whites.

**Table 2.** Components of Net Worth by Racial and Ethnic Groups: Adults ages 18 and older in New York City, June 2024. Source: New York City Racial Wealth and Health Survey

| Table 2. Components of Net Worth by Racial and Ethnic Groups: Adults ages 18 and older in New York City, June 2024 |  |  |  |  |  |  |  |  |  |  |  |  |
| --- | --- | --- | --- | --- | --- | --- | --- | --- | --- | --- | --- | --- |
| Source: New York City Racial Wealth and Health Survey |  |  |  |  |  |  |  |  |  |  |  |  |
|  |  | Puerto Rican | Dominican | Other Latino | African American, Non-Latino | Caribbean or West Indian, Non-Latino | Other Black, Non-Latino | Chinese, Non-Latino | Other Asian or Pacific Islander, Non-Latino | American Indian, Native, First Nations, Indigenous Peoples of the Americas, or Alaska Native, Non-Latino | Multi-race or some other race (including Middle Eastern/Northern African), Non-Latino | White, Non-Latino (ref group) |
| Total Number of Observations | N = 2,866 | n = 329 | n = 287 | n = 354 | n = 393 | n = 246 | n = 157 | n = 252 | n = 277 | n = 23 | n = 137 | n = 411 |
| Panel A |  |  |  |  |  |  |  |  |  |  |  |  |
| Net worth | Median Net Worth (USD) | 160 | 300 | 5,000 | 500 | 8,800 | 25 | 320,000 | 107,000 | 1,000 | 5,000 | 142,000 |
|  | Percentage in Wealth Quartile 1 (-4,362,900 USD — -4 USD) | 20.4 | 20.2 | 20.3 | 22.1 | 26.0 | 18.5 | 6.0 | 13.7 | — | 25.6 | 18.7 |
|  | Percentage in Wealth Quartile 2 (0 USD — 11,501 USD) | 45.3 | 45.3 | 34.2 | 38.4 | 26.4 | 52.9 | 15.1 | 17.0 | 56.6 | 29.2 | 13.1 |
|  | Percentage in Wealth Quartile 3 (12,000 USD — 360,060 USD) | 21.3 | 25.1 | 24.9 | 22.1 | 22.0 | 17.8 | 31.8 | 34.4 | 30.4 | 16.8 | 27.3 |
|  | Percentage in Wealth Quartile 4 (362,000 USD — 1,749,948,500 USD) | 13.1 | 9.4 | 20.6 | 17.3 | 25.6 | 10.8 | 47.2 | 35.0 | 13.0 | 28.5 | 40.9 |

| Panel B (Assets) |  |  |  |  |  |  |  |  |  |  |  |  |
| --- | --- | --- | --- | --- | --- | --- | --- | --- | --- | --- | --- | --- |
| Cash or cash equivalents<sup>a **</sup> | Percentage of households with cash or cash equivalents | 73.5 | 73.4 | 80.8 | 78.1 | 86.2 | 66.9 | 95.6 | 87.7 | 65.2 | 85.4 | 94.7 |
|  | Median amount of cash and cash equivalents | 1,000* | 2,000* | 3,000* | 1,500* | 2,200* | 1,000* | 40,000 | 20,000 | 5,000* | 5,000* | 30,000 |
| Stocks | Percentage of households owning stocks, mutual funds, or investment trust | 17.0 | 23.3 | 31.4 | 20.6 | 26.4 | 18.5 | 49.6 | 46.9 | 26.1 | 29.2 | 51.3 |
|  | Median amount | 5,000 (674 - 9,326)* | 5,000 (260 - 9,740)* | 20,000 (4,172 - 35,828)* | 20,000 (927 - 39,073)* | 40,000 (21,750 - 58,250) | 10,000 (- 31,895 - 51,895)* | 50,000 (19,292 - 80,708) | 40,000 (14,190 - 65,810) | 2,000 (-2 - 4,002)* | 40,000 (5,103 - 74,987) | 100,000 (29,722 - 147,540) |
| IRA and Private Annuity | Percentage of households having IRA or Private Annuity | 21.0 | 19.5 | 29.7 | 25.5 | 28.5 | 21.7 | 47.6 | 42.2 | 26.1 | 28.5 | 54.5 |
|  | Median amount | 20,000 (8,110 - 31,890)* | 20,000 (6,802 - 33,198)* | 18,000 (8,362 - 27,638)* | 50,000 (25,309 - 74,691) | 25,000 (13,195 - 36,805)* | 20,000 (- 2,019 - 42,019)* | 50,000 (27,911 - 72,089) | 60,000 (37,173 - 82,827) | 10,000 (- 4,112 - 24,112) | 50,000 (26,278 - 73,722) | 80,000 (34,314 - 176,862) |
| Retirement (Retirement Plans & Pensions) ** | Percentage of households with a retirement account | 28.6 | 31.4 | 33.3 | 37.2 | 45.5 | 27.4 | 36.9 | 35.7 | 21.7 | 30.7 | 39.7 |
|  | Median amount | 10,000* | 12,000* | 12,000* | 18,000 | 25,000 | 5,000* | 67,000 | 30,000 | 3,000 | 33,500 | 57,000 |
| Homeownership | Percentage of households owning a home | 18.8 | 12.5 | 24.0 | 22.1 | 31.7 | 15.3 | 49.6 | 37.2 | 8.7 | 24.8 | 34.3 |
|  | Median value of home | 520,000 (255,189 - 784,811) | 300,000 (52,874 - 547,126) | 500,000 (374,354 - 625,646) | 500,000 (400,645 - 599,355) | 600,000 (534,418 - 665,582) | 400,000 (204,213 - 595,787) | 750,000 (625,667 - 874,333) | 650,000 (538,142 - 761,858) | 400,000 (- 910,384 - 1,710,384) | 850,000 (667,229 - 1,032,771) | 700,000 (472,674 - 1,315,506) |
| Vehicle Ownership | Percentage of households owning a vehicle | 30.1 | 23.7 | 29.7 | 29.5 | 38.2 | 19.8 | 44.4 | 45.5 | 8.7 | 24.8 | 38.7 |
|  | Median value of vehicle | 13,000<br>(8,026 - 17,974) | 20,000<br>(13,999 - 26,001) | 20,000<br>(15,654 - 24,346) | 17,500<br>(10,608 - 24,392) | 20,000<br>(14,896 - 25,104) | 12,000<br>(7,556 - 16,444) | 24,000<br>(17,453 - 30,547) | 20,000<br>(13,387 - 26,613)* | 900 (-1,899 - 3,699) | 15,000<br>(4,816 - 25,184) | 15,000<br>(5,076 - 17,873) |
| Tangible Assets <sup>b **</sup> | Percent | 39.5 | 35.5 | 46.1 | 41.5 | 52.4 | 33.8 | 70.6 | 63.5 | 26.1 | 40.2 | 58.2 |
|  | Median amount (U.S. dollars) of tangible asset worth | 29,000 * | 30,000 * | 90,000 * | 40,000 * | 400,000 | 25,000 * | 500,000 | 329,500 | 2,000 | 551,000 | 400,000 |
| Liquid Assets <sup>c</sup> | Percent | 75.7 | 77.0 | 82.5 | 81.2 | 87.4 | 69.4 | 96.8 | 91.0 | 69.6 | 85.4 | 95.1 |
|  | Median amount (U.S. dollars) of liquid asset worth | 2,300 (-1,565 - 6,165)* | 6,000<br>(2,342 - 9,658)* | 14,000<br>(5,672 - 22,328)* | 8,000<br>(3,903 - 12,097)* | 11,320 (-696 - 23,336)* | 4,000 (-998 - 8,998)* | 90,100<br>(55,395 - 124,805) | 60,000<br>(36,522 - 83,478)* | 20,000 (-6,936 - 46,936)* | 10,000 (-8,794 - 28,794)* | 125,000<br>(82,556 - 194,607) |
| Total Assets | Percent | 78.1 | 79.1 | 85.0 | 81.9 | 89.4 | 71.3 | 98.4 | 93.5 | 73.9 | 86.1 | 95.6 |
|  | Median amount (U.S. dollars) of total asset worth | 15,000<br>(6,938 - 23,062)* | 17,000<br>(8,535 - 25,465)* | 40,000<br>(23,069 - 56,931)* | 26,000<br>(12,727 - 39,273)* | 60,000<br>(8,503 - 111,497)* | 11,000 (-5,755 - 27,755)* | 480,000<br>(309,132 - 650,868) | 265,090<br>(144,316 - 385,894) | 21,800 (-7,167 - 50,767)* | 75,000 (-9,128 - 159,128)* | 273,000<br>(163,536 - 439,685) |
| Panel C (Debts) |  |  |  |  |  |  |  |  |  |  |  |  |
| Credit Card Debt | Percentage of households with credit card debt | 30.7 | 30.3 | 32.8 | 37.2 | 48.0 | 21.0 | 15.1 | 28.9 | 0.0 | 38.0 | 42.8 |
|  | Median Amount of Credit Card Debt | 7,000<br>(4,586 - 9,414) | 6,000<br>(3,771 - 8,229) | 11,000<br>(7,783 - 14,217) | 7,000<br>(4,951 - 9,049) | 10,000<br>(7,152 - 12,848) | 8,500<br>(5,053 - 11,947) | 10,000<br>(5,182 - 14,818) | 12,000<br>(6,465 - 17,535) | — | 10,000<br>(5,538 - 14,462) | 9,000<br>(4,196 - 24,018) |
| Student Loans | Percentage of households with student loans | 16.4 | 13.2 | 20.6 | 18.1 | 29.7 | 16.6 | 7.9 | 18.1 | 0.0 | 19.7 | 26.3 |
|  | Median Amount of Student Loans | 20,000<br>(14,417 - 25,583)* | 30,000<br>(18,687 - 41,313) | 22,000<br>(13,837 - 30,163) | 45,000<br>(35,263 - 54,737) | 45,000<br>(30,595 - 59,405) | 46,000<br>(10,600 - 81,400) | 30,000 (-3,758 - 63,758) | 40,000<br>(23,755 - 56,245) | — | 30,000<br>(10,262 - 49,738) | 43,000<br>(25,792 - 70,208) |
| Health Care Debt | Percentage of households with health care debt | 9.1 | 8.0 | 9.9 | 12.0 | 11.8 | 7.0 | 3.6 | 5.8 | 0.0 | 10.2 | 12.2 |
|  | Median Amount of Health Care Debt | 3,000 (1,233 - 4,767) | 3,000 (-767 - 6,767) | 1,500 (-676 - 3,676) | 3,000 (1,306 - 4,694) | 5,000 (2,304 - 7,696) | 2,000 (541 - 3,459) | 10,000 (-7,853 - 27,853) | 10,000 (4,354 - 15,646) | — | 3,500 (-1,674 - 8,674) | 2,000 (2,000 - 5,970) |
| Legal Debt | Percentage of households with legal debt | 0.6 | 3.5 | 2.0 | 1.0 | 0.8 | 3.8 | 0.8 | 1.8 | 0.0 | 2.2 | 2.0 |
|  | Median Amount of Legal Debt | 3,000 (-196 - 6,196) | 3,000 (-4,061 - 10,061) | 7,000 (-33,995 - 47,995) | 4,000 (-68,292 - 76,292) | 80,000 (-21,505 - 181,505) | 5,000 (-2,971 - 12,971) | 10,000 (-1,122 - 21,122) | 20,000 (11,915 - 28,085) | — | 2,500 (-7,931 - 12,931) | 3,500 (2,070 - 40,118) |
| Installment Loans | Percentage of households with installment loans | 4.9 | 6.3 | 5.4 | 7.4 | 12.6 | 3.8 | 4.0 | 4.3 | 0.0 | 4.4 | 5.8 |
|  | Median Amount of Installment Loans | 25,000 (10,371 - 39,629) | 12,000 (4,119 - 19,881) | 10,000 (1,050 - 18,950) | 7,000 (4,930 - 9,070) | 7,500 (1,494 - 13,506) | 4,000 (-25,008 - 33,008) | 10,000 (-3,217 - 23,217) | 18,000 (1,108 - 34,892) | — | 18,000 (-26,365 - 62,365) | 11,000 (4,163 - 31,722) |
| Property Mortgage or Loan ** | Among all homeowners, percentage with mortgage debt | 54.8 | 50.0 | 57.7 | 41.4 | 68.0 | 62.5 | 46.4 | 58.3 | 100.0 | 61.8 | 61.0 |
|  | Median value of remaining principal on a mortgage or loan among all homeowners | 168,000* | 100,000 | 185,000 | 160,000 | 256,000 | 280,000 | 250,000 | 270,000 | 1,000 | 243,560 | 280,000 |
| Vehicle Debt | Among all vehicle owners, percentage with vehicle debt | 44.4 | 48.5 | 46.7 | 48.3 | 51.1 | 35.5 | 45.5 | 42.9 | 100.0 | 41.2 | 27.7 |
|  | **Median value of vehicle debt among all vehicle owners** | 10,000<br>(4,098 - 15,902) | 10,000<br>(1,563 - 18,437) | 10,000 (413 - 19,587) | 14,000<br>(7,025 - 20,975) | 15,000<br>(9,619 - 20,381) | 10,000<br>(8,876 - 11,124) | 20,000<br>(13,735 - 26,265) | 15,000<br>(8,912 - 21,088) | 8,000 (- 12,300 - 28,300) | 14,500<br>(8,073 - 20,927) | 15,000<br>(8,480 - 30,520) |
| **Secured Debt <sup>d \*\*</sup>** | **Percentage of households with secured debt** | 34.4 | 26.8 | 35.0 | 32.1 | 43.9 | 24.2 | 53.6 | 54.5 | 13.0 | 30.7 | 47.2 |
|  | **Median Amount of Secured Debt** | 3,000* | 729* | 6,290* | 2,100* | 25,000 | 10,000 | 20,000 | 15,000 | 1,300 | 30,000 | 15,000 |
| **Unsecured Debt <sup>e</sup>** | **Percentage of households with unsecured debt** | 38.0 | 38.0 | 40.1 | 42.5 | 56.1 | 30.6 | 21.8 | 41.2 | 0.0 | 49.6 | 52.3 |
|  | **Median Amount of Unsecured Debt** | 16,125<br>(10,854 - 21,396) | 16,000<br>(9,290 - 22,710) | 23,000<br>(15,333 - 30,667) | 18,000<br>(11,420 - 24,580) | 24,000<br>(16,309 - 31,691) | 16,200<br>(3,160 - 29,240) | 20,000<br>(7,961 - 32,039) | 30,000<br>(18,952 - 41,048) | — | 21,000<br>(14,555 - 27,445) | 30,000<br>(17,646 - 41,177) |
| **Any type of Debt <sup>\*\*</sup>** | **Percentage of households with any type of debt** | 55.6 | 53.3 | 58.2 | 56.5 | 71.1 | 42.0 | 61.1 | 72.6 | 13.0 | 62.8 | 77.4 |
|  | **Median Amount of Debt** | 16,000* | 14,900* | 25,000 | 20,000* | 45,000 | 18,000* | 25,000 | 33,000 | 1,300 | 29,000* | 45,000 |

<sup>a</sup> "Cash or cash equivalents" is the total amount within checking and savings accounts, money market funds, certificates of deposits, government savings bonds, and treasury bills.
<sup>b</sup> Tangible assets are physical assets that have a finite monetary value and can be quantified (i.e., houses, vehicles, and other properties one may own).
<sup>c</sup> Liquid assets are cash in hand or assets that can be readily converted to cash at or near their current market value (i.e., cash, stocks, bonds, and money market instruments).
<sup>d</sup> Secured debt is debt that have collateral, which is a specific piece of property that the lender can take back if the money is not paid back (i.e., mortgage and vehicle debt).
<sup>e</sup> Unsecured debt is debt not backed by collateral or an underlying asset (i.e. credit card debt, student loans and loans from family and friends)
\*Statistically significant when compared to White, Non-Latino reference group in quartile regression
\*\* The quantile regression did not generate 95% confidence interval of medians for these measures and that comparisons with respect to White medians were done based on 95% confidence intervals of the differences in medians.

A similar pattern is observed across all specific asset types. For instance, White respondents are 2-3 times as likely as Other Black or Puerto Rican respondents to own stock (51% vs. 21% and 17%, respectively), and more than twice as likely as Dominican respondents to have Individual Retirement Accounts (IRAs) or private annuities (55% vs. 20%, respectively). Quantile regression results confirm significant disparities in both cash reserves and retirement savings, with Puerto Rican and Dominican adults reporting significantly lower median values compared with Whites. Homeownership is highest among Chinese (50%), Other API (37%), and White (34%) respondents and lowest among Dominican (13%) and Other Black (15%) respondents (Table 2).

In terms of debts, over 70% of White, Caribbean, and Other API respondents hold debts, with median values of $45,000, $45,000, $30,000 respectively. Dominican, Puerto Rican, and Other Black respondents report lower median debt values than other groups, ranging from $14,900 to $18,000. Puerto Rican, Dominican, and African American adults had significantly lower secured and total debt relative to Whites. Chinese and Other API respondents, however, did not differ significantly from Whites in secured or total debt levels.

Chinese respondents have one of the highest percentages of secured debt ownership (54%, with a median of $20,000) while simultaneously reporting among the lowest percentages of unsecured debt (22%, with a median of $20,000) across all racial and ethnic groups. Black and Latino respondents, specifically Puerto Rican (34%), Dominican (27%), and Other Black (24%), report lower percentages of secured debt and moderate to high levels of unsecured debt, from 31% to 38%, and median unsecured debt amounts between $16,000 and $16,200. White (47%) and multiracial (31%) respondents reported high levels of both secured and unsecured debt, with 52% and 50% of households holding unsecured debt and median unsecured debt amounts of $30,000 and $21,000, respectively (Table 2).

### Health Status by Wealth and Racial/Ethnic Groups

The percentage of respondents reporting excellent or very good health rises from 36% in the lowest wealth quartile to 59% in the highest. Reports of low or no psychological distress also increase with wealth, from 70% in wealth quartile 1 to 86% in wealth quartile 4 (Table 3).

**Table 3.** Health Characteristics by Wealth Distribution: Adults ages 18 and older in New York City, June 2024. Source: New York City Racial Wealth and Health Survey

|  | Total |  | Wealth Quartile 1<br>(-4,362,900 USD — -4 USD) |  | Wealth Quartile 2<br>(0 USD — 11,501 USD) |  | Wealth Quartile 3<br>(12,000 USD — 360,060 USD) |  | Wealth Quartile 4<br>(362,000 USD — 1,749,948,500 USD) |  |
| --- | --- | --- | --- | --- | --- | --- | --- | --- | --- | --- |
|  | N = 2,866 | Column % | n = 542 | Column % (95% Confidence Interval) | n = 891 | Column % (95% Confidence Interval) | n = 716 | Column % (95% Confidence Interval) | n = 717 | Column % (95% Confidence Interval) |
| **Self-Reported Health Status** |  |  |  |  |  |  |  |  |  |  |
| Excellent or very good | 1,360 | 47.5 | 196 | 36.2 (32.1 - 40.2) | 384 | 43.1 (39.8 - 46.4) | 360 | 50.3 (46.6 - 53.9) | 420 | 58.6 (55.0 - 62.2) |
| Good | 967 | 33.7 | 194 | 35.8 (31.8 - 39.8) | 312 | 35.0 (31.9 - 38.2) | 246 | 34.4 (30.9 - 37.8) | 215 | 30.0 (26.6 - 33.3) |
| Fair or Poor | 539 | 18.8 | 152 | 28.0 (24.3 - 31.8) | 195 | 21.9 (19.2 - 24.6) | 110 | 15.4 (12.7 - 18.0) | 82 | 11.4 (9.1 - 13.8) |
| **Presence of a Chronic Disease (i.e. diabetes, hypertension, asthma) <sup>a</sup>** |  |  |  |  |  |  |  |  |  |  |
| No | 1,536 | 53.6 | 277 | 51.1 (46.9 - 55.3) | 471 | 52.9 (49.6 - 56.1) | 387 | 54.1 (50.4 - 57.7) | 401 | 55.9 (52.3 - 59.6) |
| Yes | 1,330 | 46.4 | 265 | 48.9 (44.7 - 53.1) | 420 | 47.1 (43.9 - 50.4) | 329 | 45.9 (42.3 - 49.6) | 316 | 44.1 (40.4 - 47.7) |
| **Psychological Distress** |  |  |  |  |  |  |  |  |  |  |
| Low or No Psychological Distress | 2,264 | 79.0 | 381 | 70.3 (66.4 - 74.1) | 701 | 78.7 (76.0 - 81.4) | 563 | 78.6 (75.6 - 81.6) | 619 | 86.3 (83.8 - 88.8) |
| High Psychological Distress | 602 | 21.0 | 161 | 29.7 (25.9 - 33.6) | 190 | 21.3 (18.6 - 24.0) | 153 | 21.4 (18.4 - 24.4) | 98 | 13.7 (11.2 - 16.2) |

<sup>a</sup>. This variable does not include individuals with a chronic disease diagnosis only during pregnancy (e.g., diabetes or hypertension diagnosed only during pregnancy).

The prevalence estimates of several health indicators vary across groups. More than half (59%) of White respondents report excellent or very good health compared to 40% of African Americans. African American respondents report the highest percentage with chronic diseases (60%) compared with the Multiracial (34%). Most African American and Caribbean respondents (86%) report low psychological distress, respectively, compared with over two-thirds of Multiracial respondents (69%; Supplementary Table 2).

Across all racial and ethnic groups, those in the highest wealth quartile are more likely than those in the lowest quartile to self-report excellent or very good health (Figure 1). We observe similar trends for low or no psychological distress. As wealth increases, health improves more among some racial and ethnic groups than among others. For instance, Chinese and Multiracial respondents show the most pronounced differences in self-reported excellent/very good health across wealth quartiles, increasing from 13% to 52% and 34% to 67%, respectively, while African American respondents show little variation beyond the lowest wealth quartile.

**Figure 1.**
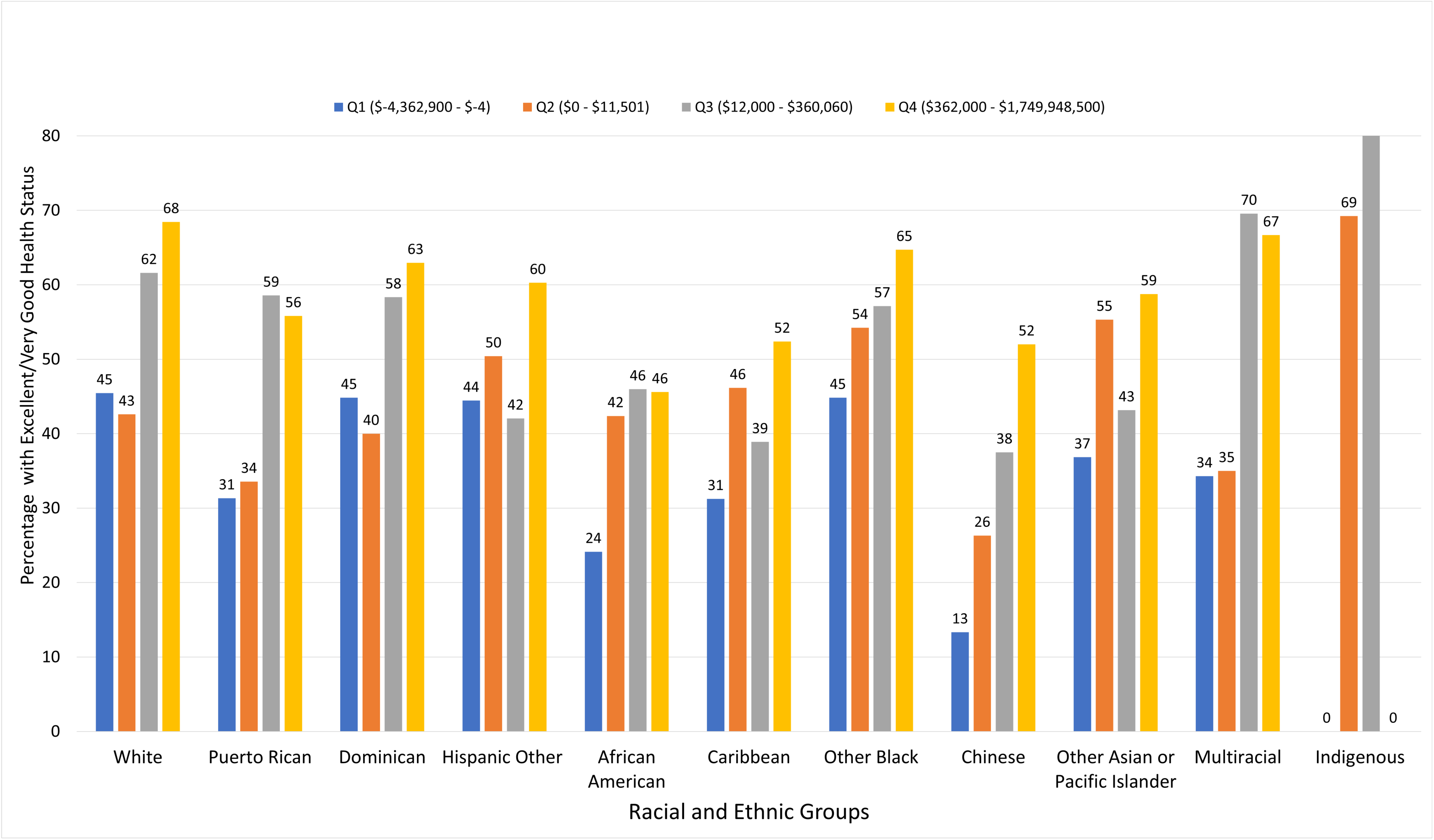
Excellent/Very Good Health Status by Wealth Quartiles and Racial and Ethnic Groups in New York City, June 2024

White respondents were more likely than every other group in the highest wealth quartile, and almost every other group in the second-highest quartile, to self-report excellent or very good health.

## Discussion

We sought to characterize racial/ethnic inequalities in health and wealth in a diverse sample of New Yorkers. Our findings show that, across racial and ethnic groups in NYC, wealth is generally associated with better health outcomes, although the pattern varies across groups. Additionally, we found poorer self-reported health among nearly all non-white racial and ethnic groups than among white respondents within the two highest wealth quartiles, but a more complicated picture in the two lower quartiles. Our findings are consistent with Color of Wealth studies conducted in other U.S. cities, highlighting persistent racial wealth inequities where Black and Latino populations consistently hold the lowest wealth levels. This pattern mirrors trends seen in cities like Chicago and Boston, where Black households similarly have near-zero wealth as in NYC [2, 24].

Our findings suggest that, within our sample, Black and Latino households in NYC hold less than 10% of the wealth that white households do, as in Los Angeles [25] and Tulsa [27]. Puerto Rican and Dominican respondents have lower wealth than other Latino respondents in NYC, as in Miami [28]. Together, these findings support existing evidence that structural forces at the national level and/or across multiple U.S. jurisdictions may have perpetuated racial wealth inequity, and solutions at multiple levels will likely be required to correct it.

Wealth provides long-term stability, offers a buffer against health emergencies, helps bridge insurance gaps, and enables access to healthier environments and high-quality care. However, racialized groups may derive fewer physical and mental health benefits from wealth due to factors like residential segregation, asset devaluation in communities of color, and discriminatory experiences in health and financial systems. Across the life course, Black adults with similar net worth to their white counterparts report worse physical and mental health, pointing to diminished “health returns” on wealth for minoritized groups. These inequities are often compounded by systemic disinvestment and neighborhood-level exposures, such as environmental toxins, policing, and food insecurity, that disproportionately affect lower-wealth communities of color.

Our study is unique in its local focus and has several strengths and limitations. We leveraged a large administrative database and intentionally oversampled specific racial and ethnic groups to produce a local picture of race, wealth, and health, reflecting NYC’s demographic diversity. However, several limitations need to be underscored. The online-only administration excluded individuals without internet access. The sampling frame was not representative of the NYC population as a whole based on the non-probability construct of the databases. This was mitigated by employing a stratified quota sampling approach to ensure adequate representation in the final sample across ancestry groups. Indigenous people were reached at a lower rate than other groups, but among those who were reached, response was the highest. However, a substantial portion of Indigenous respondents were excluded from analyses because of missing key data elements, limiting the findings for this group. Taken together, these issues may have further compromised the representativeness of the sample and increased sampling error. Additionally, while aiming to measure household net worth, the survey relied on individual self-reports of family-level financial values, and some questions unintentionally elicited responses about individual rather than household-level finances, both of which potentially introduced measurement errors. Despite these limitations, this study presents an innovative and informative approach to an understudied topic.

## Public Health Implications

A core public health function is to identify and address health hazards and root causes and remove the systemic barriers that create inequalities. NYC, arguably the most diverse and wealthiest large U.S. city, is also characterized by deep economic inequality and racial segregation. Bold public policies are needed to address wealth and health gaps [31]. Local action requires local data, and wealth should become a routinely collected indicator in public health efforts nationwide. Our findings can inform state and city commitments to evidence-based solutions to address the lasting harms of discriminatory policies and practices. Reparations programs are one such solution, with modeling studies suggesting they could help eliminate wealth and health gaps nationally [15, 21, 32]. Though narrower in scope, New York has state and city commitments to investigate, document, and address the lasting harms of years of discriminatory practices in the U.S. [33, 34]. Progressive tax reform is also needed to the racial wealth gap and to generate sustained investments in historically marginalized communities [35, 36].

Measuring wealth as a population health metric acknowledges the historical inequalities rooted in systemic and structural racism. Our study suggests that including wealth in routine public health surveillance is feasible and offers more locally relevant insights than national surveys. Our findings may shape local public policy to address the legacy of racism, reduce racial health inequities, and reduce wealth inequality in the NYC population.

## Data Availability

All data produced in the present study is not available.

## Acknowledgments

We are grateful to Emerson College for administering the survey. Specifically, Patrick Fox, Matt Taglia, Camille Mumford, and Spencer Kimball. We thank the following persons for their methodological advisement: Stephen Immerwahr and Sungwoo Lim of the NYC Health Department, Dr. Lauren Russell of the Federal Reserve Board, Dr. Raffi Garcia of Rensselaer Polytechnic Institute, Dr. Gwendolyn Wright of Duke / Cook Center, Dr. Sara Chaganti and Dr. Mike Evangelist of the Federal Reserve Bank of Boston, Dr. Laura Sullivan of the New Jersey Institute for Social Justice, Dr. Suparna Bhaskaran and Elaine Chang of the New School Institute for Race, Power, and Policy, and Dr. Chris Wimer and Schuyler Ross of Columbia University.

---

**Supplementary Table 1.**
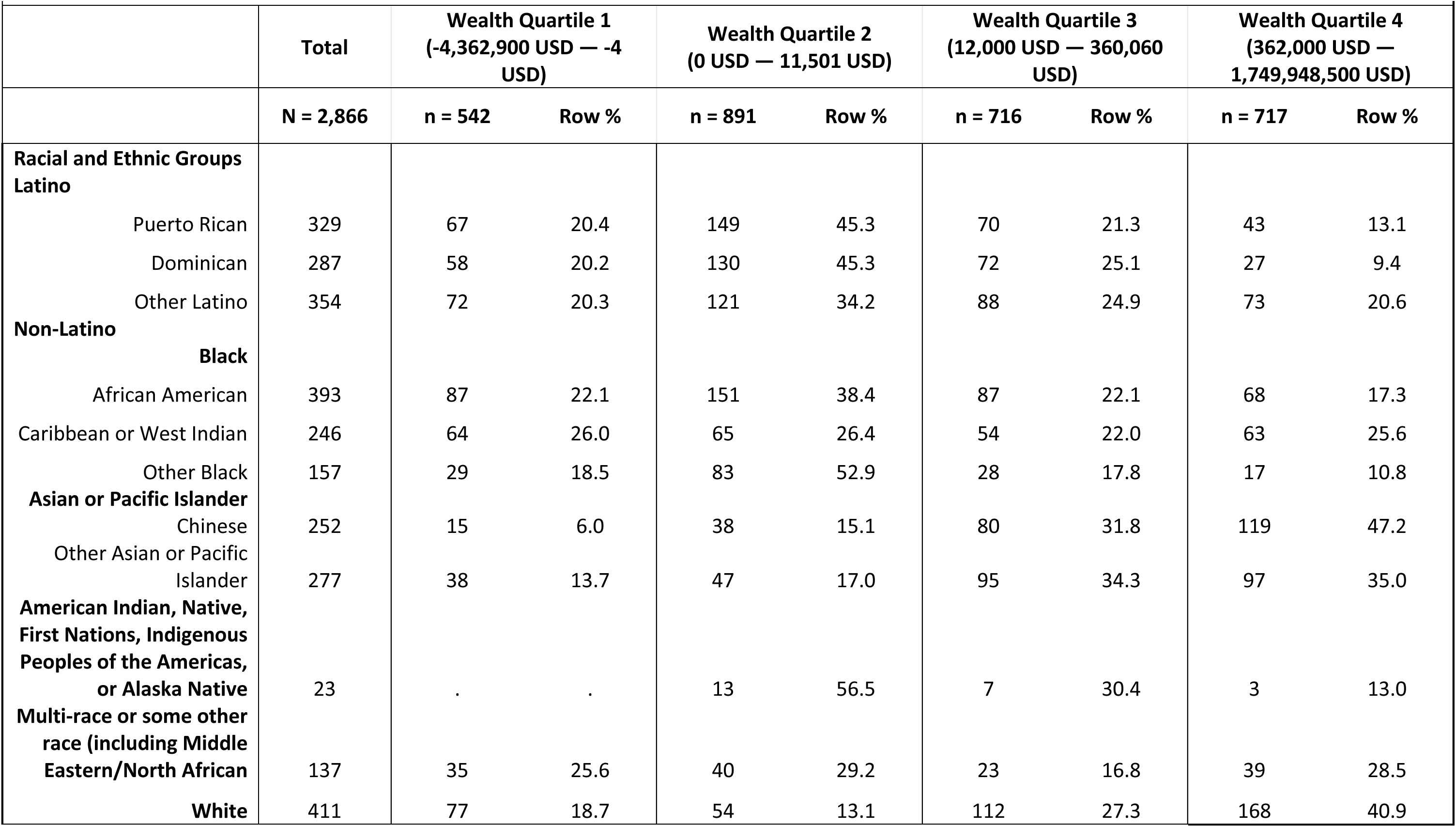
Racial and Ethnic Groups by Wealth Distribution: Adults ages 18 and older in New York City, June 2024. Source: New York City Racial Wealth and Health Survey

**Supplementary Table 2.** Sample Health Characteristics by Racial and Ethnic Groups: Adults ages 18 and older in New York City, June 2024. Source: New York City Racial Wealth and Health Survey

| Supplementary Table 2. Sample Health Characteristics by Racial and Ethnic Groups: Adults ages 18 and older in New York City, June 2024 |  |  |  |  |  |  |  |  |  |  |  |  |  |  |  |  |  |  |  |  |  |  |
| --- | --- | --- | --- | --- | --- | --- | --- | --- | --- | --- | --- | --- | --- | --- | --- | --- | --- | --- | --- | --- | --- | --- |
| Source: New York City Racial Wealth and Health Survey |  |  |  |  |  |  |  |  |  |  |  |  |  |  |  |  |  |  |  |  |  |  |
|  | Racial and Ethnic Groups |  |  |  |  |  |  |  |  |  |  |  |  |  |  |  |  |  |  |  |  |  |
|  | Puerto Rican |  | Dominican |  | Other Latino |  | African American, non-Latino |  | Caribbean or West Indian, non-Latino |  | Other Black, non-Latino |  | Chinese, non-Latino |  | Other Asian or Pacific Islander, non-Latino |  | American Indian, Native, First Nations, Indigenous Peoples of the Americas, or Alaska Native, non-Latino |  | Multi-race/MENA/Some other race, non-Latino |  | White, non-Latino |  |
| Overall Sample | n | % | n | % | n | % | n | % | n | % | n | % | n | % | n | % | n | % | n | % | n | % |
| Insurance Status |  |  |  |  |  |  |  |  |  |  |  |  |  |  |  |  |  |  |  |  |  |  |
| Insured | 313 | 95.1 | 266 | 92.7 | 329 | 92.9 | 373 | 94.9 | 231 | 93.9 | 143 | 91.1 | 240 | 95.2 | 262 | 94.6 | 20 | 87.0 | 122 | 89.1 | 387 | 94.2 |
| Uninsured | 14 | 4.3 | 9 | 3.1 | 17 | 4.8 | 11 | 2.8 | 9 | 3.7 | 3 | 1.9 | 5 | 2.0 | 10 | 3.6 | 1 | 4.4 | 7 | 5.1 | 18 | 4.4 |
| Don't know their insurance status | 2 | 0.6 | 12 | 4.2 | 8 | 2.3 | 9 | 2.3 | 6 | 2.4 | 11 | 7.0 | 7 | 2.8 | 5 | 1.8 | 2 | 8.7 | 8 | 5.8 | 6 | 1.5 |
| Self-Reported Health Status |  |  |  |  |  |  |  |  |  |  |  |  |  |  |  |  |  |  |  |  |  |  |
| Excellent/Very Good | 136 | 41.3 | 137 | 47.7 | 174 | 49.2 | 156 | 39.7 | 104 | 42.3 | 85 | 54.1 | 104 | 41.3 | 138 | 49.8 | 16 | 69.6 | 68 | 49.6 | 242 | 58.9 |
| Good | 114 | 34.7 | 91 | 31.7 | 112 | 31.6 | 162 | 41.2 | 92 | 37.4 | 46 | 29.3 | 103 | 40.9 | 91 | 32.9 | 5 | 21.7 | 45 | 32.9 | 106 | 25.8 |
| Fair/Poor | 79 | 24.0 | 59 | 20.6 | 68 | 19.2 | 75 | 19.1 | 50 | 20.3 | 26 | 16.6 | 45 | 17.9 | 48 | 17.3 | 2 | 8.7 | 24 | 17.5 | 63 | 15.3 |
| Presence of a Chronic Disease (i.e. diabetes, hypertension, asthma) |  |  |  |  |  |  |  |  |  |  |  |  |  |  |  |  |  |  |  |  |  |  |
| Yes | 186 | 56.5 | 121 | 42.2 | 165 | 46.6 | 237 | 60.3 | 121 | 49.2 | 74 | 47.1 | 105 | 41.7 | 100 | 36.1 | 10 | 43.5 | 46 | 33.6 | 165 | 40.2 |
| No | 143 | 43.5 | 166 | 57.8 | 189 | 53.4 | 156 | 39.7 | 125 | 50.8 | 83 | 52.9 | 147 | 58.3 | 177 | 63.9 | 13 | 56.5 | 91 | 66.4 | 246 | 59.9 |
| Psychological Distress |  |  |  |  |  |  |  |  |  |  |  |  |  |  |  |  |  |  |  |  |  |  |
| Low or No Psychological Distress | 243 | 73.9 | 202 | 70.4 | 266 | 75.1 | 339 | 86.3 | 212 | 86.2 | 126 | 80.3 | 208 | 82.5 | 216 | 78.0 | 16 | 69.6 | 94 | 68.6 | 342 | 83.2 |
| High Psychological Distress | 86 | 26.1 | 85 | 29.6 | 88 | 24.9 | 54 | 13.7 | 34 | 13.8 | 31 | 19.8 | 44 | 17.5 | 61 | 22.0 | 7 | 30.4 | 43 | 31.4 | 69 | 16.8 |
| Last Routine Check-Up |  |  |  |  |  |  |  |  |  |  |  |  |  |  |  |  |  |  |  |  |  |  |
| One year or less | 242 | 73.6 | 216 | 75.3 | 239 | 67.5 | 319 | 81.2 | 189 | 76.8 | 117 | 74.5 | 186 | 73.8 | 196 | 70.8 | 17 | 73.9 | 102 | 74.5 | 257 | 62.5 |
| 1-5 years ago | 63 | 19.2 | 46 | 16.0 | 77 | 21.8 | 53 | 13.5 | 39 | 15.9 | 23 | 14.7 | 49 | 19.4 | 59 | 21.3 | 1 | 4.4 | 24 | 17.5 | 114 | 27.7 |
| 5 or more years ago | 4 | 1.2 | 4 | 1.4 | 14 | 4.0 | 7 | 1.8 | 9 | 3.7 | 2 | 1.3 | 8 | 3.2 | 9 | 3.3 | . | . | 4 | 2.9 | 23 | 5.6 |
| Don't know/not sure | 12 | 3.7 | 14 | 4.9 | 16 | 4.5 | 6 | 1.5 | 6 | 2.4 | 6 | 3.8 | 4 | 1.6 | 10 | 3.6 | 4 | 17.4 | 2 | 1.5 | 9 | 2.2 |
| Never | 8 | 2.4 | 7 | 2.4 | 8 | 2.3 | 8 | 2.0 | 3 | 1.2 | 9 | 5.7 | 5 | 2.0 | 3 | 1.1 | 1 | 4.4 | 5 | 3.7 | 8 | 2.0 |
| **Unmet medical care in the past 12 months** |  |  |  |  |  |  |  |  |  |  |  |  |  |  |  |  |  |  |  |  |  |  |
| Yes, foregone medical care for a reason other than cost | 41 | 12.5 | 33 | 11.5 | 46 | 13.0 | 39 | 9.9 | 20 | 8.1 | 21 | 13.4 | 21 | 8.3 | 23 | 8.3 | 2 | 8.7 | 12 | 8.8 | 39 | 9.5 |
| Yes, foregone medical care due to cost | 51 | 15.5 | 55 | 19.2 | 69 | 19.5 | 42 | 10.7 | 27 | 11.0 | 18 | 11.5 | 19 | 7.5 | 45 | 16.3 | 2 | 8.7 | 28 | 20.4 | 61 | 14.8 |
| No, did not forego medical care | 237 | 72.0 | 199 | 69.3 | 239 | 67.5 | 312 | 79.4 | 199 | 80.9 | 118 | 75.2 | 212 | 84.1 | 209 | 75.5 | 19 | 82.6 | 97 | 70.8 | 311 | 75.7 |
| **Unmet mental health care and/or treatment in the past 12 months** |  |  |  |  |  |  |  |  |  |  |  |  |  |  |  |  |  |  |  |  |  |  |
| Yes | 77 | 23.4 | 57 | 19.9 | 60 | 17.0 | 46 | 11.7 | 24 | 9.8 | 23 | 14.7 | 34 | 13.5 | 54 | 19.5 | 6 | 26.1 | 31 | 22.6 | 80 | 19.5 |
| No | 228 | 69.3 | 194 | 67.6 | 261 | 73.7 | 316 | 80.4 | 209 | 85.0 | 113 | 72.0 | 203 | 80.6 | 197 | 71.1 | 11 | 47.8 | 94 | 68.6 | 291 | 70.8 |
| Do Not Know | 24 | 7.3 | 36 | 12.5 | 33 | 9.3 | 31 | 7.9 | 13 | 5.3 | 21 | 13.4 | 15 | 6.0 | 26 | 9.4 | 6 | 26.1 | 12 | 8.8 | 40 | 9.7 |
| **Facility visited most often for care...** |  |  |  |  |  |  |  |  |  |  |  |  |  |  |  |  |  |  |  |  |  |  |
| A doctor's private office | 142 | 43.2 | 115 | 40.1 | 158 | 44.6 | 186 | 47.3 | 134 | 54.5 | 63 | 40.1 | 182 | 72.2 | 170 | 61.4 | 8 | 34.8 | 65 | 47.5 | 268 | 65.2 |
| Community health center | 65 | 19.8 | 66 | 23.0 | 53 | 15.0 | 80 | 20.4 | 44 | 17.9 | 30 | 19.1 | 27 | 10.7 | 24 | 8.7 | 4 | 17.4 | 17 | 12.4 | 30 | 7.3 |
| A hospital outpatient clinic | 60 | 18.2 | 47 | 16.4 | 62 | 17.5 | 67 | 17.1 | 39 | 15.9 | 22 | 14.0 | 23 | 9.1 | 38 | 13.7 | 4 | 17.4 | 19 | 13.9 | 48 | 11.7 |
| An urgent care center, such as CityMD or ProHealth | 34 | 10.3 | 24 | 8.4 | 32 | 9.0 | 34 | 8.7 | 14 | 5.7 | 14 | 8.9 | 6 | 2.4 | 21 | 7.6 | . | . | 15 | 11.0 | 40 | 9.7 |
| A hospital emergency room | 11 | 3.3 | 11 | 3.8 | 13 | 3.7 | 9 | 2.3 | 6 | 2.4 | 6 | 3.8 | 2 | 0.8 | 2 | 0.7 | 3 | 13.0 | 6 | 4.4 | 3 | 0.7 |
| A retail clinic such as CVS Minute-Clinic | 2 | 0.6 | 3 | 1.1 | 4 | 1.1 | 3 | 0.8 | 2 | 0.8 | 2 | 1.3 | . | . | 3 | 1.1 | . | . | . | . | 1 | 0.2 |
| Other: Some other place, no usual place, or don't know where they receive care | 15 | 4.6 | 21 | 7.3 | 32 | 9.0 | 14 | 3.6 | 7 | 2.9 | 20 | 12.7 | 12 | 4.8 | 19 | 6.9 | 4 | 17.4 | 15 | 11.0 | 21 | 5.1 |
| **Work limitations** |  |  |  |  |  |  |  |  |  |  |  |  |  |  |  |  |  |  |  |  |  |  |
| Yes | 105 | 31.9 | 61 | 21.3 | 84 | 23.7 | 108 | 27.5 | 41 | 16.7 | 37 | 23.6 | 34 | 13.5 | 50 | 18.1 | 2 | 8.7 | 42 | 30.7 | 86 | 20.9 |
| No | 208 | 63.2 | 212 | 73.9 | 256 | 72.3 | 271 | 69.0 | 195 | 79.3 | 113 | 72.0 | 205 | 81.4 | 213 | 76.9 | 18 | 78.3 | 87 | 63.5 | 314 | 76.4 |
| Don't Know | 16 | 4.9 | 14 | 4.9 | 14 | 4.0 | 14 | 3.6 | 10 | 4.1 | 7 | 4.5 | 13 | 5.2 | 14 | 5.1 | 3 | 13.0 | 8 | 5.8 | 11 | 2.7 |

